# COVID-19 in Cambodia, January 2020–June 2022: a success story

**DOI:** 10.1101/2022.12.24.22283920

**Authors:** Srean Chhim, Grace Marie Ku, Sovathiro Mao, Willem van de Put, Wim Van Damme, Por Ir, Chhea Chhorvann, Vandine Or

**Affiliations:** National Institute of Public Health, Phnom Penh, Cambodia; Institute of Tropical Medicine, Antwerp, Belgium; Vrije Universiteit Brussel, Belgium; Ministry of Health, Phnom Penh, Cambodia

**Keywords:** COVID-19, SARS-CoV-2, Cambodia, Response, Lessons learned

## Abstract

As a member state of the International Health Regulation 2005, Cambodia has been continuously strengthening its capacity to respond to health emergencies and prevent the international spread of diseases. Despite this, Cambodia’s capacity to prevent, detect, and rapidly respond to public health threats remained limited at the onset of the pandemic. This paper describes epidemiological phases, response phases, strategy, and lessons learned in Cambodia between 27 January 2020 and 30 June 2022. We classified epidemiological phases in Cambodia into three phases, in which Cambodia responded using eight measures: *(1) detect, isolate/quarantine; (2) face coverings, hand hygiene, and physical distancing measures; (3) risk communication and community engagement; (4) school closures; (5) border closures; (6) public event and gathering cancellation; (7) vaccination;* and *(8) lockdown*. The measures corresponded to six strategies: *(1) setting up and managing a new response system, (2) containing the spread with early response, (3) strengthening the identification of cases and contacts, (4) strengthening care for COVID-19 patients, (5) boosting vaccination coverage*, and *(6) supporting disadvantaged group*s. Finally, ten lessons were learned for future health emergency responses. Findings suggest that Cambodia successfully contained the spread of SARS-CoV-2 in the first year and quickly attained high vaccine coverage by the second year of the response. The core of this success was the strong political will and high level of cooperation from the public. However, Cambodia needs to further improve its infrastructure for quarantining and isolating cases and close contacts and laboratory capacity for future health emergencies.

**SUMMARY BOX:**

- COVID-19 spread globally, but how the pandemic played out in each country depended on various factors, including government responses and the general public’s adherence to COVID-19 measures.
- Early response—*Early detection, Early isolation, Early tracing, Early treatment, and Early education*—is the core of successful SARS-CoV-2 containment.
- Achieving high vaccination coverage quickly leads to a decline in the number of deaths and to eventual full re-opening of the country.
- Responding to the pandemic requires decisive leadership and good governance, that refers to decisions being made quickly, in a timely manner, and without delay.
- High level of cooperation from the public is a fundamental factor for success in containing the spread in the early phase, and the massively successful vaccination campaign in the later stage.

## INTRODUCTION

Starting in early 2020, COVID-19 spread globally. However, how the epidemic played out in each country depended on various factors, including the government responses and the general public’s adherence to COVID-19 measures.[1]

Cambodia’s health system has significantly improved in the last three decades contributing to a rise in life expectancy.[2] However, its readiness in responding to a health emergency was still limited at the onset of the pandemic. In 2014, the World Health Organization (WHO) estimated that Cambodia had only 0.2 physicians per 1,000 population.[3, 4] The ratio of hospital beds was also low at 0.9 per 1,000 population.[4, 5] There was limited infrastructure for isolation of patients with highly infectious diseases.

As a member state of the International Health Regulation (IHR) 2005, Cambodia committed to implementing the IHR’s framework and continuously strengthening its capacity to respond to health emergencies, including outbreaks. In spite of this, the Joint External Evaluation by the IHR—a process of evaluating a country’s capacity to prevent, detect, and rapidly respond to public health threats—indicated that Cambodia had limited capacity in 2016.[6]

The COVID-19 response in Cambodia has not been well-documented in scientific publications. While several papers described the early phases of COVID-19 responses including non-pharmaceutical interventions (NPIs), many other important aspects, such as rapid scale up of laboratory capacity and vaccination, were overlooked.[7, 8]

Our paper describes the epidemiological phases, response phases, strategy, and lessons learned in Cambodia between 27 January 2020 (when the first case was detected) and 30 June 2022, several months after Cambodia’s full re-opening.

## METHODOLOGY

We analysed secondary data and reviewed relevant literature and documents to describe the COVID-19 epidemiology, responses, and strategies between 27 January 2020 and 30 June 2022 in Cambodia.

### Data for descriptive epidemiology

#### Tests, cases, and deaths

The Inter-ministerial COVID-19 Response Committee provided the data of COVID-19 tests, and we used publicly available data from WHO’s COVID-19 dashboard for cases and deaths.[9] The tests, cases, and deaths recorded between 27 January 2020 and 30 June 2022. The COVID-19 data is under the governance and stewardship of the Inter-ministerial COVID-19 Response Committee, with Cambodia’s Communicable Disease Control Department (CCDC) being the central body responsible for collection and storage. The data were aggregated and reported to WHO as part of the IHR 2005’s commitment. The data were collected from all laboratories in Cambodia, using a laboratory form (see supplementary file 1). The completed forms were entered into each laboratory database and exported to a CSV or Excel file and submitted to the CCDC. At the CCDC, these files were checked for standardisation, and imported into the National COVID-19 Database.

### Data for response and themes

#### Relevant events

We documented relevant events to draw a more comprehensive picture of the COVID-19 response in Cambodia. Our data team recorded all the policies and guidelines issued by the government, and triangulated these documents with other documents (e.g., WHO website updates, situation reports). The information we collected included dates of border closures, school closures, cancellation/prohibition of public events and gatherings, enforcement of facemask usage in public areas, and lockdowns.

#### Vaccination data

The National COVID-19 Vaccination Committee provided vaccination data. These data included the number of vaccine doses that can be stratified by vaccine brand. It also included vaccination coverage data: the number of vaccinated individuals, and the number of doses administered (e.g., one dose/incomplete immunisation, two doses/completed immunisation, first booster doses, second booster doses, and third booster doses).

## RESULTS AND DISCUSSION

### EPIDEMIOLOGICAL PHASES

Figure 1 presents the daily number of COVID-19 cases, and PCR tests in Cambodia. In Cambodia, we identified three significant phases of the pandemic: containment phase, mitigation phase, and full re-opening phase.

**Figure 1.**
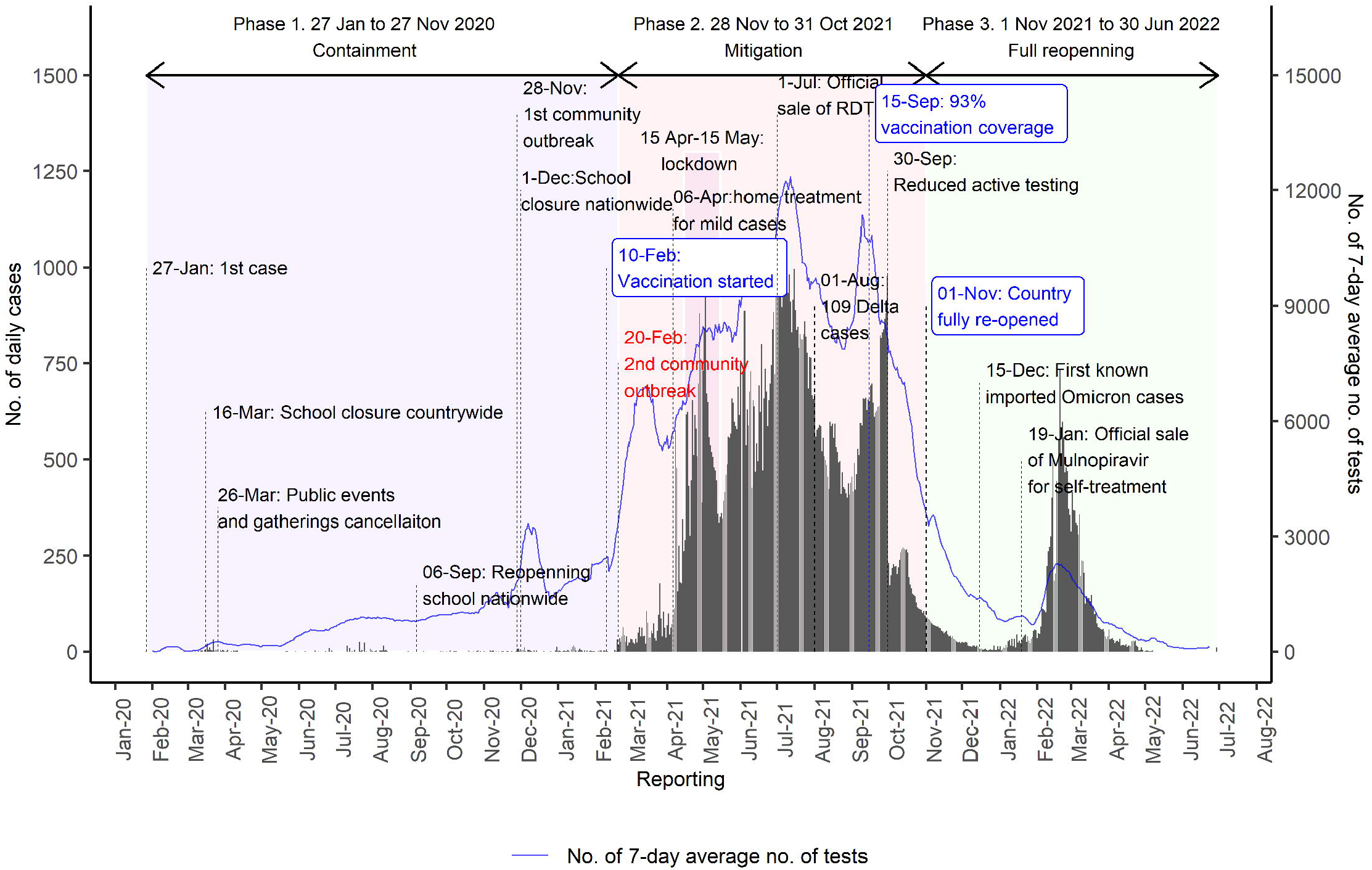
COVID-19 daily cases, tests, and deaths in Cambodia, 2020–2022. Note: Data is based on the official data from the MOH.

#### Phase 1 (27 January to 19 February 2021): containment phase

In Phase 1, COVID-19 was well-contained. Cambodia had only 484 cases, mainly imported and no deaths (Figure 1 and Table 1).

**Table 1.** Reported COVID-19 cases and deaths by phase in Cambodia, 27 January 2020 to 30 June 2022

| Phase of COVID-19 outbreak | Theme | All cases | Death |
| --- | --- | --- | --- |
| **Phase 1. 27 January 2020 to 19 February 2021:** | Containment with NPIs | 484 | 0 |
| *27 January–30 April 2020* | Initial response<br>• Intensive contact tracing<br>• Quarantine/isolation<br>• Border restriction | 122 | 0 |
| *1 May–31 October 2020* | Relaxing after long period of low cases | 169 | 0 |
| *1 November 2020–19 February 2021* | Intensifying response for larger scale outbreak | 193 | 0 |
| **Phase 2. 20 February 2021 to 31 October 2021** | Large community outbreak and vaccination<br>• Vaccine rolling out since 10 February 2021<br>• City lockdown<br>• Travel restrictions<br>• School closures<br>• Home treatment/isolation<br>• Government cash transfers to disadvantaged groups | 118,129 | 2,781 |
| **Phase 3. 1 November 2021 to 30 June 2022** | Full re-opening<br>• Achieved goal: 10 million population vaccinated<br>• Continue with 3 DOs and 3 DON'Ts<br>• Continue to provide booster dose and make it routine<br>• Continue to support the disadvantaged groups | 17,669 | 275 |

Abbreviations: NPIs, Non-pharmaceutical interventions

#### Phase 2 (20 February to 31 October 2021): mitigation phase

In Phase 2, a large community outbreak occurred that lasted for 252 days with 118,129 cases (Figure 1 and Table 1). A total of 2,781 COVID-19 deaths were reported (Figure 2 and Table 1).

**Figure 2.**
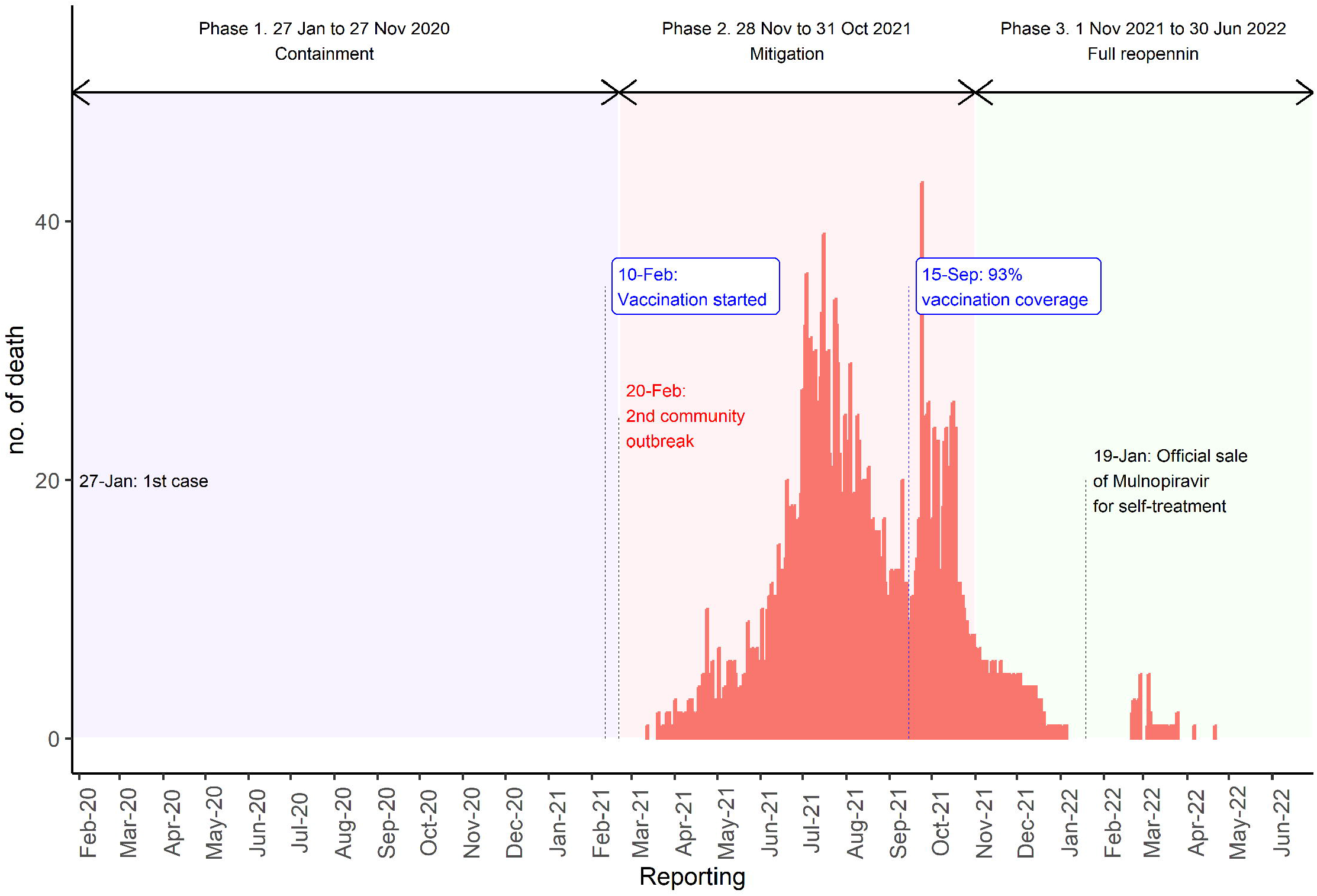
COVID-19 death trends in Cambodia, 2021–2022. Note: Data is based on the official data from the MOH.

#### Phase 3 (1 November 2021 to 30 June 2022): full re-opening

Phase 3 had 17,669 cases (Figure 1 and Table 1). In this phase, 275 deaths were reported.

### RESPONSE PHASES

The government implemented eight significant measures during phases 1 and 2: *(1) detect, isolate/quarantine; (2) face coverings, hand hygiene, and physical distancing measures; (3) risk communication and community engagement; (4) school closures; (5) border closures; (6) public event and gathering cancellation; (7) vaccination; and (8) lockdown* (Table 2). Each of these measures was implemented quickly when the need for such intervention was seen to be warranted. The first six measures were implemented in phase 1, and measures 7 and 8 were added in phase 2.

**Table 2:**
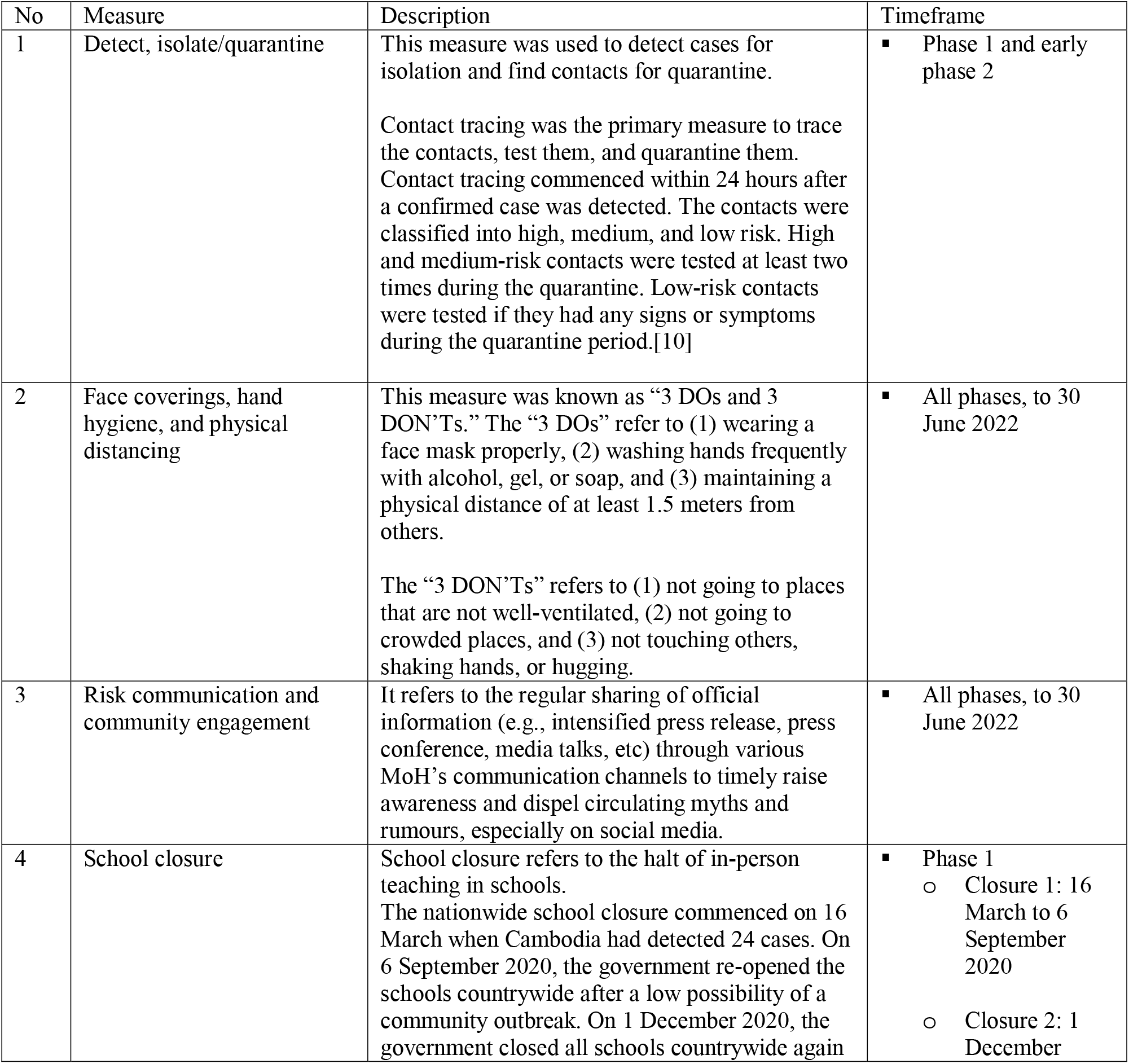
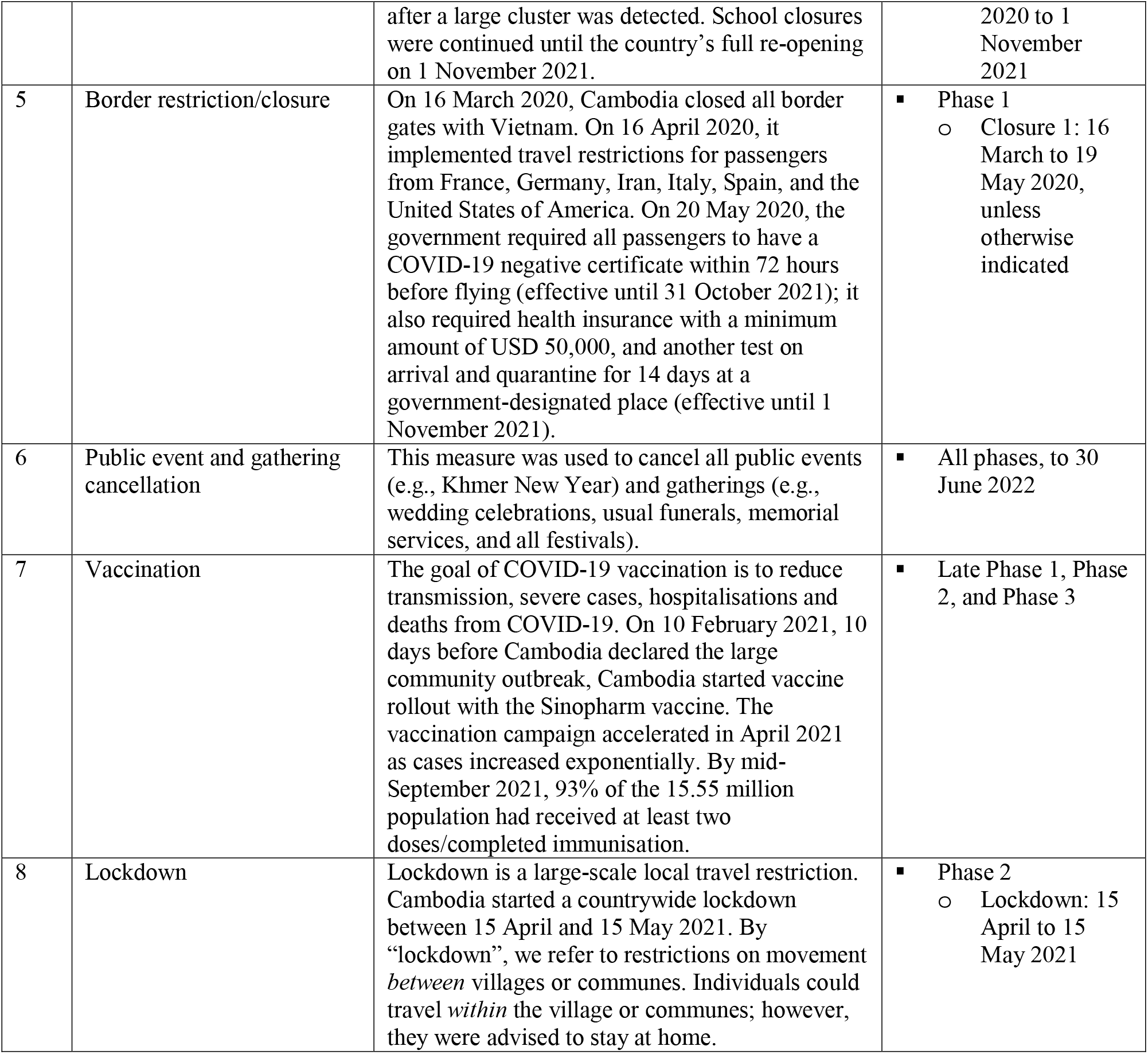
Description of the eight measures used in Cambodia in response to the COVID-19 outbreak

#### Phase 1. 27 January 2020 to 19 February 2021: containment with NPIs

After detecting the first case, on 30 January 2020, Cambodia started implementing mandatory temperature screenings for all travellers entering Cambodia at all points of entry (airports and international borders). In March and April 2020, Cambodia detected several clusters of imported cases that were not identified through the temperature screening system, but through notifications from the WHO’s IHR channel and event-based surveillance (screening local and international news). From these primary cases, the virus spread locally.

To respond to these clusters, intensive contact tracing was undertaken to trace, test and quarantine the contacts. This strategy was an essential component of “*Measure 1: detect, isolate/quarantine”*. Contact tracing commenced within 24 hours after a case was detected. All contacts were quarantined at quarantine centres or homes for 14 days from the last date of exposure.

Cambodia also implemented *Measure 2: face covering, hand hygiene, and physical distancing* since it detected its first case. The government strongly advised people to take such measures for self-protection and compliance by the population was very high.

The government, through the Ministry of Health (MoH) also implemented *Measure 3: risk communication and community engagement* from the onset. Regular sharing of official information through various MoH communication channels helped to dispel circulating myths, false information, and rumours, especially on social media.[11] The MoH reiterated simple messages to the public, known as *“3 DOs and 3 DON’Ts”*. The “3 DOs” refer to (1) wearing a face mask properly, (2) frequent hand disinfection using alcohol, gel-based formulations, or soap, and (3) maintaining a physical distance of at least 1.5 meters from others. The “3 DON’Ts” refer to (1) not going to places that are not well-ventilated, (2) not going to crowded places, and (3) not touching others, shaking hands, or hugging. These nationwide communication campaigns stimulated cooperation from the public. They were launched by the MoH through the “Sub-Committee on Education, Training, and Public Communication of the Inter-ministerial Committee to Fight COVID-19” with the theme *“Acting together to stop COVID-19 transmission”*.

On 16 March 2020, when the country’s total number of detected cases reached 24, *Measure 4: school closures* were imposed nationwide. As 23 of the cases (95.8%) were imported, Cambodia also implemented *Measure 5: border closures*. Cambodia first closed all border gates with Vietnam before full closure with other countries. On 26 March 2020, when Cambodia reached 90 cases, Measure *6: cancellation of all public events* (e.g., Khmer New Year) *and gatherings* (e.g., wedding celebrations, usual funeral memorial services, all festivals) were imposed. Travel restrictions for passengers from France, Germany, Iran, Italy, Spain, and the United States of America were implemented on 16 April 2020.

By 20 May 2020, following the increasing trend of cases, travel restrictions were modified and the government required all passengers to have a COVID-19 negative certificate within 72 hours before flying, health insurance with a minimum cover of USD 50,000, another test on arrival, and quarantine for 14 days at a government-designated place.

After a few months with low cases, the government considered Cambodia had a low-risk of transmission outbreak and decided to re-open all schools nationwide on 6 September 2020.

On 3 November 2020, the Hungarian Foreign Minister came on a two-day visit to Cambodia and met several hundred government officials including the Prime Minister, as well as students, and teachers, on different occasions. On 6 November 2020, he tested positive for SARS-CoV-2 on his return in Bangkok, Thailand. This became locally known as the “3 November Event”. Everyone who attended these gatherings and their contacts (over 2,000 people) were tested and quarantined at home for 14 days since the last date of exposure. Four cases were detected as part of this cluster. It was unclear who the source of infection was, as the Hungarian Foreign Minister tested negative within 72 hours before flying to Cambodia. The “3 November Event” prompted the government to tighten all measures due to its high potential for community transmission. On 23 November 2020, the government announced the closure of the “3 November Event” after no more cases were detected.

On 28 November 2020, Cambodia announced its first community outbreak after a woman hospitalised with respiratory symptoms and later tested positive for SARS-CoV-2, locally known as the “28 November community outbreak”. Six of her family members also tested positive. They had no history of travel abroad. However, she and her family visited several provinces in Cambodia and shopping centres for “Black Friday”. As a response, massive testing and quarantine were implemented. All individuals who were at sites where one or more of the confirmed cases had visited were tested. Certain commercial businesses (e.g., AEON supermarket) were closed for 14 days. On 1 December 2020, the government re-instated nationwide school closures for in-person classes and ordered online classes to be administered instead. This remained in effect until the country’s full re-opening. Within the span of one month, over 20,000 people were tested and home-quarantined as part of this cluster. Forty-one cases (no deaths) were detected from this event. On 29 December 2020, the government announced the closure of the “28 November community outbreak”.

#### Phase 2. 20 February 2021 to 31 October 2021: mitigation with vaccination campaign

Ten days before this phase, on 10 February 2021, Cambodia started *Measure 7: rolling out COVID-19 vaccination* with the Sinopharm vaccine after China donated one million doses of the said vaccine.

On 19 February 2021, two foreign women escaped the quarantine hotels to visit a night club while waiting for test results. Their results were positive for SARS-CoV-2. It triggered a large-scale testing of the night club’s visitors. Over 20 cases were identified. On the same date, the laboratory of the National Institute of Public Health also detected several cases among those who had requested pre-travel testing.

On 20 February 2021, Cambodia announced a second community outbreak, locally known as the “20 February community outbreak”.

*Measure 1: trace, isolate/quarantine* was intensified. All individuals in the buildings or sites where one or more confirmed cases had visited were tested. Around 8,000 tests were performed per day, which was the full capacity of Cambodia’s laboratories at that time. However, as the cases increased exponentially (Figure 1), the other five measures were also intensified.

*Measure 8: lockdown* was implemented covering the period between 15 April and 15 May 2021. Despite these more stringent measures, cases continued to rise (Figure 1).

During the same period, from April 2021, the vaccination campaign was *sped up* and ran without interruption. On average, 50,000 doses were provided daily and quickly achieved vaccination coverage of 93% of the total population within five months (n = 14,485,634 out of 15,552,211) by 15 September 2021.

After achieving such high vaccination coverage, the government stopped active surveillance, but monitored two indicators—severe cases and deaths—to ensure that the COVID-19 situation remained under control.

#### Phase 3. 1 November 2021 to 30 June 2022: full re-opening

Without any unusual increase in severe cases and deaths for 30 days in a row, the government decided to lift all compulsory containment measures from 1 November 2021. However, the government still advised people to continue with the “3 DOs and 3 DON’Ts”, provided booster doses routinely, and continued to support the disadvantaged groups.

### STRATEGY

We identified six themes from the COVID-19 responses in Cambodia.

#### Theme 1: Setting up and managing a new response system

Cambodia learned quickly and adapted to the dynamics of the disease. When the number of COVID-19 cases started to increase from March 2020, the Prime Minister led the national response. He created and chaired the National COVID-19 Response Committee. This committee was composed of high-level committees to give advice on the various aspects of the country’s COVID-19 response. The high-level committees included 25 Provincial COVID-19 Committees, each chaired by the provincial governors, a High-level Ministry of Health Task Force to mobilise resources and the Technical Working Group led by Cambodia’s CCDC with technical support from governmental and non-governmental stakeholders to provide technical advice on the COVID-19 emergency response.

#### Theme 2: Containing the spread with early response

As described previously, six measures were implemented to contain the spread of the virus. These measures had a background-strategy, the so-called “5 Es”— Early Detection, Early Isolation, Early Tracing, Early Treatment, and Early Education.

#### Theme 3: Strengthening the identification of cases and contacts

Cambodia trained new rapid response teams (RRTs), increasing from around 2,000 to over 5,000 by the end of phase 1. The capacities of existing laboratories were increased and 18 new laboratories in Phnom Penh and 11 provinces were set up. Testing capacity increased from 500 PCR tests per day in April 2020, to around 15,000 PCR tests per day in July 2021.

#### Theme 4: Strengthening care for COVID-19 patients

To reduce the case-fatality rate, Cambodia had to strengthen the capacity of its health care system.[12] Cambodia addressed this issue by training many doctors and nurses to manage COVID-19 cases.

During phase 1, Cambodia trained public healthcare providers from over 1,300 public healthcare facilities, including health centres, to treat and care for COVID-19 patients. In addition, it trained doctors-and nurses-in-training to treat and care for COVID-19 patients.

School buildings, wedding buildings, and abandoned hotels were temporarily repurposed as isolation and treatment centres.

#### Theme 5: Boosting vaccination coverage

In April 2021, when the cases seemed unstoppable, Cambodia boosted the vaccination rate to build herd immunity.

After basic vaccination with the Sinovac, Sinopharm, Moderna, AstraZeneca, or Janssen vaccines, adult booster doses were given with AstraZeneca and Pfizer vaccines. Children received the Sinovac and/or Pfizer (paediatric) vaccines and booster doses.

A total of 54.06 million doses of COVID-19 vaccine were obtained (Table 3). The majority of the vaccines was Sinovac (63.7%), followed by Sinopharm (15.0%), Pfizer (adults and paediatric; 10.0%), AstraZeneca (7.8%), Covishield (1.2%), and the remainder were Janssen and Moderna (2.3%).

**Table 3.** Vaccine doses obtained by the Cambodian government till 05 February 2023

| Type of vaccine | Doses received | Percent |
| --- | --- | --- |
| Sinovac | 34,424,800 | 63.7 |
| Sinopharm | 8,100,000 | 15.0 |
| Pfizer | 4,489,290 | 8.3 |
| Pfizer Paediatric | 923,990 | 1.7 |
| AstraZeneca | 4,224,040 | 7.8 |
| Janssen | 1,060,100 | 2.0 |
| Covishield | 649,000 | 1.2 |
| Moderna | 188,160 | 0.3 |
| Total | 54,059,380 | 100.0 |

Abbreviation: ASEAN, Association of Southeast Asian Nations

#### Theme 6: Supporting disadvantaged groups

Lockdown negatively impacts those in disadvantaged groups, particularly, daily-wage earners (e.g., garment workers, Tuk-tuk (motorcycle) and taxi drivers, construction workers, and street vendors).[13] The government and local and international development partners strived to respond in a timely manner to help the disadvantaged groups.[13] Initially, delivering food supplies was too slow and challenging due to the absence of a food distribution infrastructure specifically for the poor.[8, 13, 14] Nevertheless, between May 2020 and May 2021, the government provided social assistance through the COVID-19 Cash Transfer Programme to 700,000 low-income households, equivalent to more than 2.8 million individuals. By June 2021, each household had received an average of USD 366 in nine instalments.[15]

### LESSONS LEARNT

From Cambodia’s COVID-19 response, we highlight ten lessons learnt.

#### 1. COVID-19 response requires decisive leadership and good governance

The COVID-19 pandemic is a “wicked problem” [16] and many decisions were made amidst uncertainties. Leadership is a core part of governance, referring to the government’s ability to provide guidance, plan, and make appropriate modifications to the laws or regulations at all levels and sectors to enable an effective response. We learned that responding to the pandemic requires decisive leadership, referring to decisions being taken fast, timely, and without delay. The decisive leadership must be supported by the good governance to effectively implement the newly introduced policies or guideline. With this characteristic, Cambodia responded promptly, despite limited pre-pandemic capacity. Literature suggests that decisive leadership led to success in responding to COVID-19 at the early phase in China, New Zealand, Taiwan, and South Korea.[17-21]

#### 2. Laboratory capacity determine the response speed

The country should be able to conduct tests quickly without having to ship samples overseas. Laboratory capacity limit the speed of the response, meaning delay in releasing of results also delays response, impacting community transmission and deaths due to COVID-19. A rapid scale-up of laboratory capacity was essential. An example of high mortality and excess deaths due to delays in providing COVID-19 results can be seen in Ecuador.[22] Taiwan and Singapore learned from the SARS response in 2000s and understood well the need to extend laboratory capacity and took early actions that may have significantly contributed to its success in containing the virus spread in the early stage and overall mortality rate over time.[23] In Cambodia, the Rapid Respond Team (RRT) demands an immediate laboratory result (within 24 hours), so that they can decide in the field. Cambodia managed to scale up its laboratory rapidly to response to the need of response team and dynamic of the disease.

#### 3. Risk communication and community engagement could fight the infodemics, but must be consistent

Effective risk communication and community engagement have been critical to educating people to receive reliable information, gain trust and strengthen engagement with the public.[24] Fighting the infodemic must be intense. It is important to agree that only one person or group shares the COVID-19 information to make it consistent and avoid public confusion.

#### 4. Contact tracing worked well in a small-scale response and stopped working when the community outbreak started

Contact tracing was not only to find the cases, isolate them, and find their contacts, but it also provided real-time information, which was necessary for the response teams to decide where to impose additional restrictions. However, it only works in settings where cooperation from the public is high. Moreover, it only works before community outbreak occurs. Contact tracing should be stopped when the evidence of community outbreak is confirmed to save resources for a mitigation plan.

#### 5. School closures caused class disruption, but it saved elderly lives

Schools are a perfect environment for transmission, and a risk for elderlies (aged 50 and over) who usually live with their children and grandchildren. However, school closures were highly controversial due to their effects on children’s learning. Cambodia implemented early school closures and empty schools were used as quarantine centers. This measure may have been necessary to help Cambodia successfully contain the virus for more than a year and prevent deaths from COVID-19.

#### 6. Face coverings, hand hygiene, physical distancing, border restriction/closures, and public event and gathering cancellation were not controversial in Cambodia

People in Cambodia tended to easily accept face coverings, hand hygiene, and physical distancing, border restriction/closure, and public event and gathering cancellation. The cooperation was high.

#### 7. Lockdown must accompany support to disadvantaged groups

Lockdown is a controversial measure for many settings as it affects the livelihoods of many individuals.[25] Supporting the needs of low-income families was essential to reducing the social burden of COVID-19. The slow support was due to the limited existing registration system of people who need support. However, the government managed to provide food and cash support to around 700,000 households or 2.8 million individuals.[15] From this experience, it is important to register and regularly update those who need support or so-called IDPoor in Cambodia.[26]

#### 8. Vaccination gave Cambodia a ticket to re-open the country

As a low-income country, Cambodia was unique in achieving high vaccination coverage quickly. Upon achieving a 93% vaccination coverage in mid-September 2021, a marked decline in the number of deaths was noted by October 2021, approximately one month later. Cambodia was then able to fully re-open the country.

#### 9. Cooperation from the public was the necessary factor

From the measures described above, adherence and compliance from the general public is required.[7] The government called it solidarity. This compliance is a fundamental factor for success in containing the spread in the early phase, and the very successful vaccination campaign in the later stage.

#### 10. The background-strategy of 5Es was central for success

The background strategy of the “5 Es” was behind the intensified implementation of each measure. In responding to the pandemic, every action must be timely to avoid running behind the transmission.

## CONCLUSION

Despite limited levels of preparedness prior to the pandemic, Cambodia successfully contained the spread of SARS-CoV-2 in the first year and quickly attained high vaccine coverage by the second year of the response. The core of this success was the strong political will and high cooperation from the public. However, Cambodia needs to improve its infrastructure in order to quarantine contacts isolate cases and increase its laboratory capacity to better prepare for future health emergencies. This review is among the few that presents the COVID-19 epidemiology and response, and highlights lessons learned in Cambodia for organising and investing in future responses.

This paper has not explicitly covered a few important aspects such as good governance, resource mobilization, and multilateral cooperation (e.g., role of authorities and other ministries).

Additionally, the COVID-19 data presented in this paper may be highly under-reported and may not reflect the actual trends due to limited testing capacity and changing surveillance strategies.

## Data Availability

WHO online database (Cambodia: WHO Coronavirus Disease (COVID-19) Dashboard With Vaccination Data | WHO Coronavirus (COVID-19) Dashboard With Vaccination Data)

https://covid19.who.int/region/wpro/country/kh

## ACKNOWLEDGMENTS

We would like to thank all the Inter-ministerial COVID-19 Response Committee and Cambodia’s Communicable Disease Control Department (CCDC) for providing data.

## AUTHOR CONTRIBUTIONS

SC, GK, SM, WP, WD, PI, CC, and VO designed the study; SC analyzed data; SC, GK, SM, WP, WD, PI, CC, and VO wrote the manuscript.

## ETHICS APPROVAL

Not applicable.

## DATA AVAILABILITY STATEMENT

Data is publicly available. It can be accessed via the WHO online database (<u>Cambodia: WHO Coronavirus Disease (COVID-19) Dashboard With Vaccination Data | WHO Coronavirus (COVID-19) Dashboard With Vaccination Data</u>).

