## Supplementary file 1 for "COVID-19 in Cambodia, January 2020–June 2022: a success story" (from another version of this preprint: version 1, 2022-12-31)

# ទំរង់ស្នើសុំធ្វើពិសោធន៍
### Laboratory Request Form

កំណែថ្ងៃទី ៣០ មិថុនា ២០២០
Updated on 30 June 2020

| | | | |
|---|---|---|---|
| គ្រឹះស្ថានសុខាភិបាល:<br>*Health Facility :* | | កាលបរិច្ឆេទ<br>*Date/日期* | |
| ឈ្មោះអ្នកបំពេញទំរង់:<br>*Completed by :* | | លេខទូរស័ព្ទ<br>*Telephone/電話* | |
| បំណងនៃការធ្វើតេស្ត<br>*Reasons for testing*<br>*測試目的* | ☐សង្ស័យ Suspect *懷疑* ☐អ្នកប៉ះពាល Contact *接觸* ☐តាមដាន Follow-up *複查* ☐ រលាកសួត Pneumonia *肺炎* ☐ បុគ្គលិកពេទ្យ HCW *医护人员* ☐ ផ្សេងទៀត Other________________ *其他* | | |
| បើជាអ្នកប៉ះពាល់<br>*if contact /如果联系* | ឈ្មោះអ្នកជំងឺ______________________<br>Contact with/*患者姓名* | ទំនាក់ទំនងជាមួយអ្នកជំងឺ_________________<br>Relationship with case/*与接觸者关系* | |
| ***ព័ត៌មានអ្នកជំងឺ/Patient information/患者信息*** | | | |
| ឈ្មោះអ្នកជំងឺ<br>*Patient Name/患者姓名* | | លេខសំគាល់<br>*Patient ID* | |
| ភេទ<br>*Sex /性别* | ☐ ប្រុស *Male /男* ☐ ស្រី *Female / 女* | អាយុ<br>*Age /年龄* | |
| សញ្ជាតិ<br>*Nationality /国籍* | | លេខទូរស័ព្ទ<br>*Telephone/電話* | |
| អាស័យដ្ឋាន<br>*Address /地址* | កន្លែងស្នាក់នៅ…………………………………………………………… *Residence 住所* លេខផ្ទះ………… *House 家* លេខផ្លូវ……… *Street 路号*<br>ភូមិ……………………… *Village 乡村* ឃុំ………………………… *Commune 公社* ស្រុក……………………… *District 区* ខេត្ត……………… *Province 省* | | |
| រោគសញ្ញាគ្លីនិក<br>*Clinical Symptoms*<br>*临床症状* | ☐គ្រុនក្តៅ *Fever 发烧* ☐ក្អក *Cough 咳嗽* ☐ហៀរសំបោរ *Runny Nose 流鼻涕* ☐ឈឺបំពង់ក *Sore Throat 喉咙痛* ☐ពិបាកដកដង្ហើម *Difficulty Breathing 呼吸困难* ☐គ្មាន *No symptoms 没有症状* | | |
| ថ្ងៃចេញរោគសញ្ញា<br>*Date of onset /发病日期* | | ធ្លាប់កើតជំងឺកូវីដ-១៩ឬទេ ?<br>*History of COVID-19 positive?*<br>*你曾经有过新冠肺炎病毒吗？* | ☐ទេ *No 没有* ☐មាន *Yes 有* ថ្ងៃតេស្ត: *Test Date: 检测日期* |
| ប្រវត្តិធ្វើដំណើរ<br>*Travel history/旅行历史* | ឈ្មោះខេត្ត/ប្រទេស<br>*Province/Country 国家名称* | ថ្ងៃមកដល់<br>*Date of arrival 到达日期* | |
| | លិខិតឆ្លងដែនលេខ<br>*Passport No /护照号* | លេខកៅអី<br>*Seat No/ 座号* | |
| ***ផ្នែកមន្ទីរពិសោធន៍/Laboratory*** | | | |
| ទីកន្លែងប្រមូលវត្ថុវិភាគ<br>*Place of collection* | | ថ្ងៃយកវត្ថុវិភាគ<br>*Date of collection* | |
| ប្រភេទវត្ថុវិភាគ<br>*Type of specimen* | ☐ ច្រមុះ *Nasopharyngeal* ☐ បំពង់ក *Oropharyngeal* ☐ ផ្សេងទៀត/*Others* ............................. | មន្ទីរពិសោធន៍<br>Laboratory | ☐ IPC ☐ NIPH |
| វត្ថុវិភាគលើកទី(គូសរង្វង់)<br>*N° of Sample (circle)* | 1 2 3 4 5 6 7 8 9 10 | | |
| ឈ្មោះអ្នកប្រមូល<br>*Sample collector* | | លេខទូរស័ព្ទ<br>*Telephone* | |

ហត្ថលេខា / *Signature:* ......................................................................................
